# Experiences of the COVID-19 pandemic: cross-sectional analysis of risk perceptions and mental health in a student cohort

**DOI:** 10.1101/2020.12.21.20248467

**Authors:** Ru Jia, Holly Knight, Holly Blake, Dame Jessica Corner, Chris Denning, Jonathan Ball, Kirsty Bolton, Joanne R Morling, Carol Coupland, Grazziela Figueredo, David Ed Morris, Patrick Tighe, Armando Villalon, Kieran Ayling, Kavita Vedhara

**Author notes:** Author to whom all correspondence should be addressed: Professor Kavita Vedhara, Division of Primary Care, University of Nottingham, University Park, Nottingham, NG7 2RD, UK. **Author contributions** All authors designed the study. RJ analysed the data and RJ, HK and KV wrote the first draft. All authors provided critical revisions. All authors read and approved the submitted manuscript. As corresponding author, KV attests that all listed authors meet authorship criteria and that no others meeting the criteria have been omitted. **Declaration of interest** We declare no competing interests. **Role of funding** The study was funded by the Medical Research Council (MRC), which had no further role in the study design, data collection, analysis, and interpretation, and preparation and submission of the report. JRM receives salary support from a Medical Research Council Clinician Scientist Fellowship [grant number MR/P008348/1].

## Abstract

**Objective:** This study examined the COVID-19 risk perceptions and mental health of university students on returning to campus in the midst of the COVID-19 pandemic.

**Methods:** An online survey was completed during the first four weeks of the academic year (October 2020) by 897 university students. The survey included demographics and measures of experiences of COVID-19 testing, self-isolation, shielding, perceived risk, mental health and indices capturing related psychological responses to the pandemic.

**Results:** We observed higher levels of depression and anxiety, but not stress, in students compared with pre-pandemic normative data, but lower than levels reported earlier in the pandemic in other similar cohorts. Depression, anxiety and stress were independently associated with greater loneliness and reduced positive mood. Greater worry about COVID-19 was also independently associated with anxiety and stress. Female students and those with pre-existing mental health disorders were at greatest risk of poor mental health outcomes.

**Conclusion:** Although students perceived themselves at only moderate risk of COVID-19, the prevalence of depression and anxiety among university students should remain a concern. Universities should provide adequate support for students’ mental health during term-time. Interventions to reduced loneliness and worry, and improve mood, may benefit students’ overall mental well-being.

## Introduction

The COVID-19 pandemic has unquestionably disrupted lives across the world. Increasing evidence has reported poorer mental health associated with the lockdown, especially among young people.[1-3], Despite concerns regarding risk of infection,[4] universities across the United Kingdom (UK) reopened for the autumn term in 2020. Within weeks, over 60 institutions reported COVID-19 cases and thousands of students were required to self-isolate. It became clear that in addition to an increased risk of infection, students had mental health difficulties associated with the challenges of the pandemic.[5] Although evidence from earlier in the pandemic suggested that young adults and students were at greater risk of mental health difficulties,[2, 3, 6] there has been little focus on students’ experiences while present at university. Office for National Statistics (ONS) surveyed 1,295 students from three UK universities and showed that between 12 and 18 October 2020 about half (49%) of the students reported poorer wellbeing and mental health than before the autumn term and 91% perceived COVID-19 as a risk to their physical or mental health.[6]However, it is less clear which students were at greatest risk of mental health difficulties and the psychological characteristics associated with poorer mental health outcomes. According to stress and coping theory we would not expect effects on mental health to be uniform but to vary according to an individual’s cognitive appraisal (e.g. perceived risk) and available psychosocial resources.[7]

Here we sought to add to our understanding of the effects of the pandemic on university students at the start of the academic year during the second surge of COVID-19 in the UK. We report cross-sectional findings regarding the relationship between perceived risk of COVID-19 on mental health; and consider groups who may be most at risk of mental health difficulties and the potential modifiable influences that could be the focus of future interventions.

## Materials and Methods

### Recruitment

Ethical approval was received from the University of Nottingham Faculty of Health Sciences Research Ethics Committee (FMHS 96-0920). Participants were not compensated for their participation. Students enrolled at the University of Nottingham (UK) aged 18 or over were invited to participate via recruitment emails and through a social media campaign. Recruitment took place between October 5^th^ 2020 and November 1^st^ 2020.

### Procedures

Participants provided informed consent online before completing the online survey (JISC Online Surveys). The survey included items assessing demographics and experiences of COVID-19 testing, self-isolation, shielding, and measures of perceived risk, positive mood, loneliness, COVID-19 worry (supplementary appendix Table S1) depression (Patient Health Questionnaire (PHQ-9, α= 0.90)), anxiety (Generalized Anxiety Disorder Scale (GAD- 7, α= 0.92)), and stress (Perceived Stress Scale (PSS-4, α= 0.70)).

## Results

In total, 897 students were recruited. Four participants were excluded due to being <18 years. Sixty-two per cent of the cohort was female (n=556) with an average age of 20.7 years (SD = 3.4) and predominantly consisted of undergraduates (n=789, 88%).

In terms of COVID-19 experiences, the average perceived risk of getting COVID-19 was moderate (mean=6.1, SD=2.5, range =1-10). Forty-three per cent (n=383) reported having had a National Health Service (NHS) COVID-19 test and/or an asymptomatic test administered at the University of Nottingham, among whom 29.2% (n=112) had received a positive result; 64.5 % (n=247) had received a negative result. Most participants (n=772, 84%) reported that they knew someone who had (or thought they had) COVID-19. More than half of the participants (51%, n=461) had experienced a period of self-isolation and 15% (n=142) reported shielding during the pandemic. Detailed characteristics of the cohort and COVID-19 experiences are presented in Table 1. Table 2 reveals higher mean values for depression and anxiety, compared with previously published population norms among young adults.[8, 9] Analysis according to clinical cut-offs indicated 28% and 21% met criteria for depression and anxiety respectively (supplementary appendix Table S2).[10] Mean scores for anxiety, depression, and stress were higher among students who had undertaken a NHS COVID-19 test but no difference was found between those who tested positive and negative (supplementary appendix Table S3). Univariable regression analysis showed that higher perceived risk was significantly associated with greater depression (B=0.07, CI: 0.03, 0.10, p<0.001), anxiety (B=0.08, CI: 0.05, 0.12, p<0.001), and stress (B=0.14, CI: 0.06, 0.22, p<0.001). However, when adding in demographic and psychological factors to the model, the effect of perceived risk was no longer significant. Multivariable regression analyses showed that being female and having pre-existing mental health disorders were independently associated with greater depression, anxiety, and stress. Greater loneliness and reduced positive mood were also independently associated with greater depression, anxiety, and stress; whereas worry about COVID-19 was only associated with anxiety and stress (supplementary appendix Table S4-S6).

**Table 1:** Demographics and COVID-19 experiences in the cohort (n=893)

|  | Whole sample |
| --- | --- |
| **Mean age** | 20.7 (3.4) |
| **Ethnicity** |  |
| White British | 509 (66%) |
| BAME background | 303 (34%) |
| **International students** | 197 (22%) |
| **Term-time living** |  |
| Private accommodation (alone) | 56 (6%) |
| Private accommodation (with family) | 40 (4%) |
| Private accommodation (with others) | 527 (59%) |
| Halls of residence | 270 (30%) |
| **Year of study** |  |
| 1<sup>st</sup> year Undergraduate | 233 (26%) |
| 2<sup>nd</sup> year Undergraduate | 167 (19%) |
| 3<sup>rd</sup> year Undergraduate | 260 (29%) |
| 4<sup>th</sup> Year Undergraduate | 106 (12%) |
| 5<sup>th</sup> year and above Undergraduate | 23 (3%) |
| Postgraduate | 94 (11%) |
| Other | 10 (1%) |
| **Existing physical health issue** | 83 (9%) |
| **Experience of COVID-19** |  |
| Experienced COVID-19 symptoms | 232 (26%) |
| Received NHS COVID-19 test | 334 (38%) |
| Tested positive | 111 (33%) |
| Tested negative | 199 (60%) |
| Unsure at time of report | 24 (7%) |
| Received asymptomatic COVID-19 test | 74 (8.3%) |
| Tested positive | 7 (9.5%) |
| Tested negative | 62 (83.8%) |
| Unsure at time of report | 4 (5.4%) |
| Prefer not to say | 1 (1.4%) |
| Self-isolation behaviour | 459 (51%) |
| Shielding behaviour | 142 (16%) |
| Know someone who have had (or think they have had) COVID-19 | 749 (84%) |

**Table 2:**
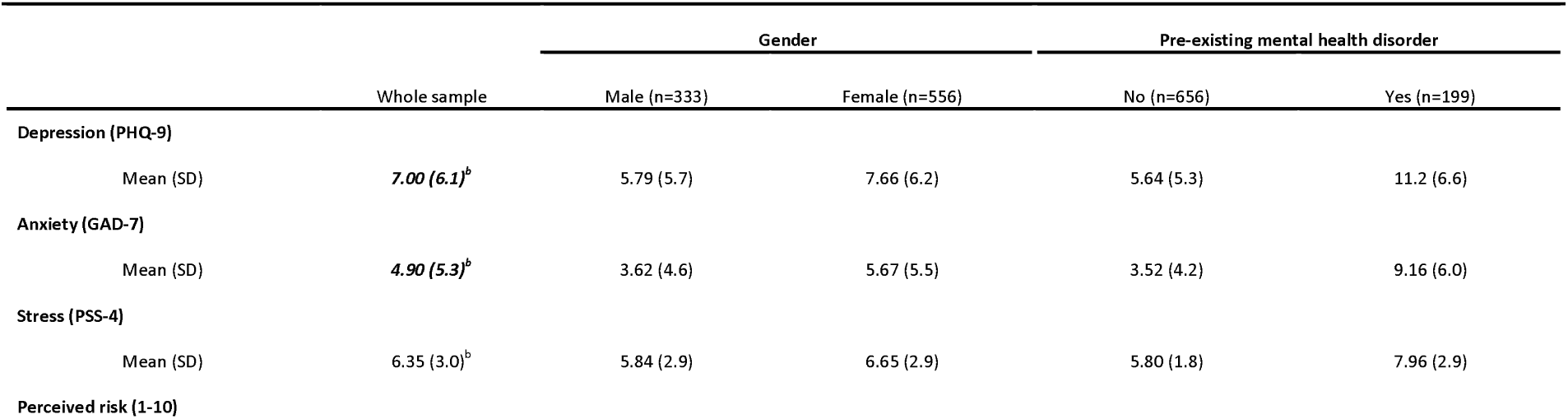
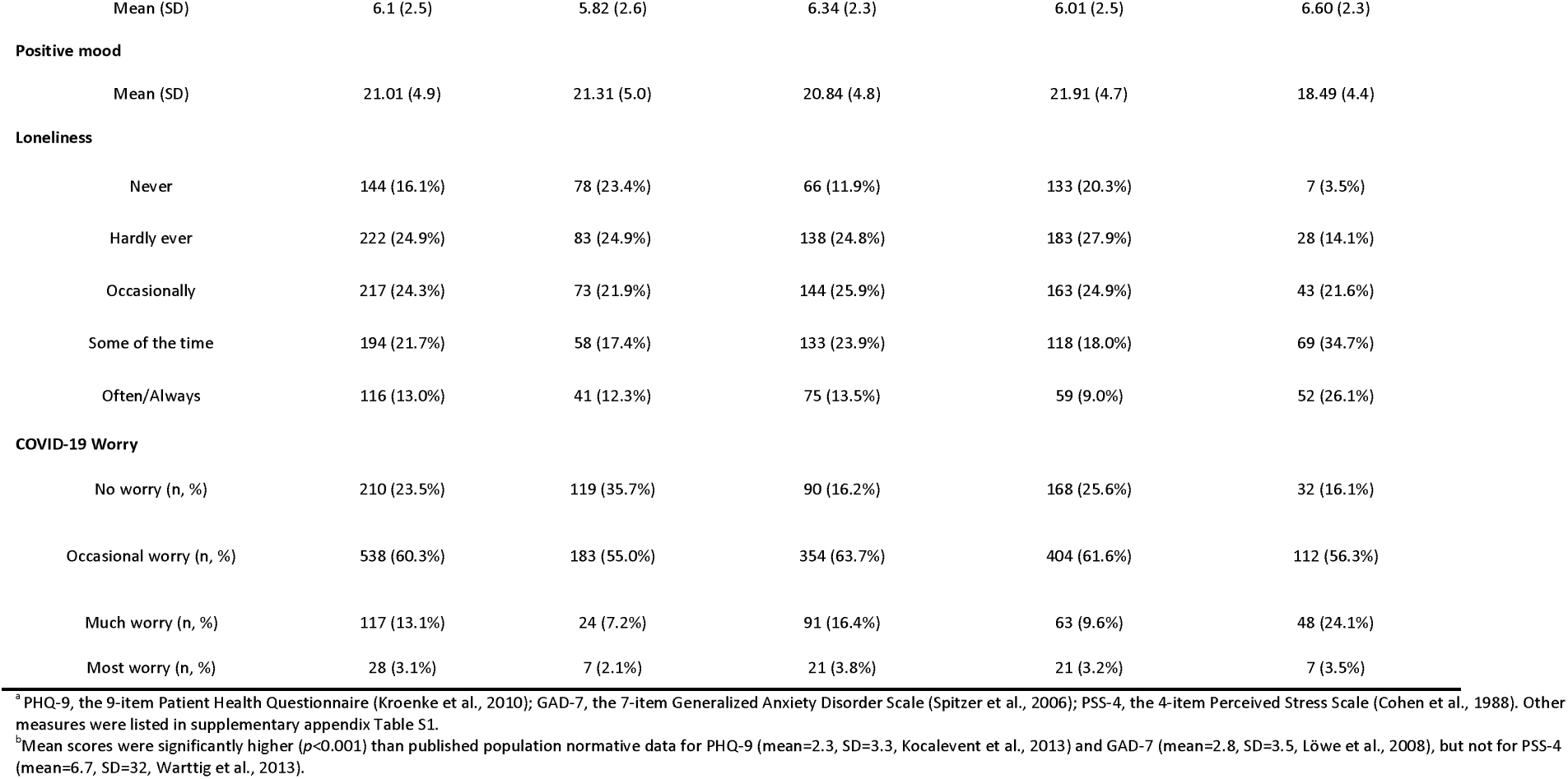
Mean scores for all measures in the cohort (n=893)^a^

## Conclusions

Our findings from a university student cohort indicated that during the first four weeks of autumn term during the second surge of COVID-19 in the UK, students perceived themselves at moderate risk of COVID-19. They also reported levels of depression and anxiety that significantly exceeded pre-pandemic norms, but these levels compared favourably with levels of anxiety and depression reported by young adults earlier in the pandemic.[1]

Several issues worthy of discussion emerge from these findings. The first concerns our mental health findings. Academic concerns, difficulty with concentration, restricted social interaction, and disrupted lifestyle resulting from remote learning and assessment caused by the pandemic may all have contributed to these poor mental health outcomes.[2, 6] However, these challenges may have become more predictable and manageable at the time of survey, and this may have led students to appraise the situation as less stressful than earlier in the pandemic, thus resulting in the attenuated distress observed in this cohort.[7] It is also possible that students perceived more available resources to cope with the stressful situation.[7] Although the majority of teaching was delivered online at the time of the research, most facilities at the participating university (including classrooms, libraries, cafes, sports facilities etc.) remained accessible. The campus environment might, therefore, have provided the context and opportunities for supporting positive mental wellbeing, despite the increased risk of COVID-19 infection.

Second, we identified demographic and psychological factors associated with increased risk of depression, anxiety, and stress. For the former, female students and those with pre-existing mental health disorders were more likely to report higher levels of depression, anxiety, and stress. This is consistent with findings from other cohorts.[2, 3, 11] For the latter, we observed associations between adverse mental health outcomes and greater loneliness and worry, and reduced positive mood with large proportion of the variance explained (24%-31%), in addition to the effect of gender and mental health history (10%-22%). These findings suggest future interventions to investigate whether reduced loneliness and worry, and improve mood, can benefit students’ overall mental well-being.

We also acknowledge several limitations. First, the cross-sectional design and absence of pre-pandemic mental health data for the same cohort prevent us from examining causal relationships. Second, our participants were from a single institution in the UK with a high proportion of female and undergraduate students, which limits the generalisability of our findings. Nonetheless, our data do indicate that the mental health of students returning to University compared favourably with the prevalence of mental health difficulties seen in a comparable age group earlier in the pandemic. However, with the means scores still substantially exceeding established norms in this population, the prevalence of depression and anxiety should remain a concern. Having students safely returning to universities requires adequate support for students’ mental health. Robust social support from the universities and wider community, with safety measures, should be offered to help students cope with loneliness and improve their mood during term-time. Finally, universities should be aware of the likely increase in the demand for counselling services, particularly amongst those with previous mental health concerns.

## Data Availability

Data will be deposited in the University of Nottingham data archive. Access to this dataset will be embargoed for a period of 12 months in order for us to complete further analysis of the dataset. After that it may be shared with the consent of the Chief Investigator.

