## Supplementary Appendix for "Experiences of the COVID-19 pandemic: cross-sectional analysis of risk perceptions and mental health in a student cohort"

Supplementary Online Appendix

**Table S1 Explanatory factors considered in the analysis**

|  | **Question/scale** | **Response(s)** |
| --- | --- | --- |
| **Non-modifiable factors** |  |  |
| Gender* | What was your gender at birth? | Male |
|  |  | Female |
|  |  | Other |
|  |  | Prefer not to say |
| Age | How old are you? | ·· |
| Ethnicity* | What is your ethnicity | White – British, Irish, other |
|  |  | Asian/Asian British – Indian, Pakistani, Bangladeshi, other |
|  |  | Black/Black British – Caribbean, African, other |
|  |  | Chinese/Chinese British |
|  |  | Mixed race – White and Black/Black British |
|  |  | Middle Eastern/Middle Eastern British – Arab, Turkish, other |
|  |  | Mixed race – other |
|  |  | Other ethnic group |
|  |  | Prefer not to say |
| Pre-existing mental disorder* | Do you have a history of anxiety, depression or any other mental health issue for which you have received treatment in the past? | Yes |
|  |  | No |
|  |  | Prefer not to say |
| **Modifiable factors** |  |  |
| *Perceived risk of COVID-19* | On a scale of 1-10, what do you believe your risk of getting COVID-19 is? | 1 (I don’t think I will get it) - 10 (I know I will most certainly get it) |
| *Positive mood^‡^* | In the past 2 weeks, I have felt Positive. | 1=Very rarely or never/ 2=Rarely/ 3=Sometimes/ 4=Often/ 5=Very often or always |
|  | In the past 2 weeks, I have felt Good. | 1=Very rarely or never/ 2=Rarely/ 3=Sometimes/ 4=Often/ 5=Very often or always |
|  | In the past 2 weeks, I have felt Pleasant. | 1=Very rarely or never/ 2=Rarely/ 3=Sometimes/ 4=Often/ 5=Very often or always |
|  | In the past 2 weeks, I have felt Happy. | 1=Very rarely or never/ 2=Rarely/ 3=Sometimes/ 4=Often/ 5=Very often or always |
|  | In the past 2 weeks, I have felt Joyful. | 1=Very rarely or never/ 2=Rarely/ 3=Sometimes/ 4=Often/ 5=Very often or always |
|  | In the past 2 weeks, I have felt Contented. | 1=Very rarely or never/ 2=Rarely/ 3=Sometimes/ 4=Often/ 5=Very often or always |
| *Loneliness* | How often have you felt lonely over the past 2 weeks? | Never |
|  |  | Hardly ever |
|  |  | Occasionally |
|  |  | Some of the time |
|  |  | Often/Always |
| *COVID-19 worry* | Please read the following statements carefully and then select the one which best describe how you have felt over the past 2 weeks. | I do not worry about getting COVID-19. |
|  |  | I occasionally worry about getting COVID-19. |
|  |  | I spend much of my time worrying about getting COVID-19. |
|  |  | I spend most of my time worrying about getting COVID-19. |

*Gender, ethnicity, and pre-existing mental disorder were treated as binary variables in all analyses: gender (male, female), ethnicity (white British, non-white British), pre-existing mental disorder (yes, no).

^†^ The factors in *Italic* were hypothesised to be associated with an increased risk of adverse mental health outcomes, apart from key-worker status where evidence exists that some key-worker roles are also associated with an increased risk of adverse COVID-19 outcomes. All other factors were hypothesised to be associated with an increased risk of contracting COVID-19 and/or poorer disease outcomes.

^‡^Positive mood was measured using the positive items from SPANE (Diener et al., 2010): Scale of Positive and Negative Experience (α=0.94).

**Mental health status among students**

Table S2 presents the categorisation of participants according to established cut-offs for depression and anxiety. This shows that 57% of the cohort reported symptoms of depression and 40% of the cohort reported symptoms of anxiety. More females reported symptoms of depression or anxiety than males. Over 82% of people who reported having a history of mental health issues reported symptoms of depression and 72% reported symptoms of anxiety. Table S4 presents the mean scores for all three mental health outcomes by testing status. Students who have had an NHS COVID-19 test reported higher levels of depression, anxiety, and stress, whilst no difference was found between student who have received positive and negative result.

**Table S2: Prevalence of depressive and anxiety cases^a^**

|  | **Categories** | **Whole sample** | **Male** | **Female** | **Mental health issue history** |
| --- | --- | --- | --- | --- | --- |
|  |  | **n (%)** | **n (%)** | **n (%)** | **n (%)** |
| **Depression (PHQ-9**^b^**)** | *No-Minimal Depression (0-4)* | 386 (43.2) | 173 (52.0) | 213 (38.3) | 35 (17.6) |
|  | *Mild Depression (5-9)* | 253 (28.3) | 90 (27.0) | 161 (29.0) | 55 (27.6) |
|  | *Moderate Depression (10-14)* | 146 (16.4) | 44 (13.2) | 102 (18.4) | 51 (25.6) |
|  | *Moderately Severe Depression (15-19)* | 66 (7.4) | 13 (3.9) | 53 (9.5) | 35 (17.6) |
|  | *Severe Depression (20-27)* | 42 (4.7) | 13 (3.9) | 27 (4.9) | 23 (11.6) |
|  | *Non-cases^c^ (0-10)* | 639 (71.6) | 263 (79.0) | 374 (67.3) | 90 (45.2) |
|  | *Cases^c^ (11-27)* | 254 (28.4) | 70 (21.0) | 182 (32.7) | 109 (54.8) |
| **Anxiety (GAD-7**^b^**)** | *No-Minimal Anxiety (0-4)* | 539 (60.4) | 237 (71.2) | 300 (54.0) | 56 (28.1) |
|  | *Mild Anxiety (5-9)* | 195 (21.8) | 60 (18.0) | 134 (24.1) | 56 (28.1) |
|  | *Moderate Anxiety (10-14)* | 94 (10.5) | 19 (5.7) | 74 (13.3) | 45 (22.6) |
|  | *Severe Anxiety (15-21)* | 65 (7.3) | 17 (5.1) | 48 (8.6) | 42 (21.1) |
|  | *Non-cases^c^ (0-8)* | 709 (79.4) | 290 (87.1) | 417 (75.0) | 104 (52.3) |
|  | *Cases^c^ (9-21)* | 184 (20.6) | 43 (12.9) | 139 (25.0) | 95 (47.7) |

^a^ Cut-offs for categories in line with published guidelines for PHQ-9 (Kroenke et al., 2010) and GAD-7 (Spitzer et al., 2006).

^b^ PHQ-9, the 9-item Patient Health Questionnaire; GAD-7, the 7-item Generalized Anxiety Disorder Scale.

*^c^* “case” is defined as a score greater than or equal to 10, at which level someone would qualify for high intensity psychological support in the NHS.

**Table S3: Mean scores for depression, anxiety, and stress by testing status**

|  | **Having had a NHS COVID-19 test** | | **NHS COVID-19 test result** | |
| --- | --- | --- | --- | --- |
|  | Yes (n=334) | No (n=559) | Positive (n=111) | Negative (n=199) |
| **Depression (PHQ-9)** |  |  |  |  |
| Mean (SD) | 7.65 (0.3)* | 6.59 (0.3) | 8.20 (0.6) | 7.24 (0.4) |
| **Anxiety (GAD-7) ^a^** |  |  |  |  |
| Mean (SD) | 5.55 (0.3)** | 4.52 (0.2) | 6.10 (0.6) | 5.26 (0.4) |
| **Stress (PSS-4)** |  |  |  |  |
| Mean (SD) | 6.69 (0.2)** | 6.16 (0.1) | 6.84 (0.3) | 6.70 (0.2) |

*** *p*<0.001, ** *p*<0.01, * *p*<0.05

Factors associated with mental health outcomes among students

Table S4-S6 present results from regression analyses examining both non-modifiable and modifiable factors associated with depression, anxiety and stress scores in the cohort. In univariable regression analyses, perceived risk was significantly associated with depression, anxiety, and stress. However, this effect was no longer significant when other factors were introduced to the models. Being female, having previous mental health issues were independently significantly associated with higher levels of depression, lower positive mood and greater loneliness were significantly associated with higher levels of depression, anxiety, and stress. Greater worry about COVID-19 was also associated with greater anxiety and stress. These factors together accounted for 46% of the variance in depression scores, 46% of the variance in anxiety scores, and 41% in stress scores.

**Table S4 Regression model showing associations between explanatory variables and depression scores**

|  | **B** | **95% CI Lower** | **95% CI Upper** | ***β*** | ***p*** |
| --- | --- | --- | --- | --- | --- |
| **PHQ-9 (depression) Total Score^a^** |  |  |  |  |  |
| Step 1 |  |  |  |  |  |
| Perceived Risk of COVID-19 (per unit) | 0.07 | 0.03 | 0.10 | 0.13 | 0.00*** |
| **Adjusted R^2^=0.02, n=893** | | | | | |
| Step 2 |  |  |  |  |  |
| Perceive Risk of COVID-19 (per unit) | 0.05 | 0.01 | 0.08 | 0.09 | 0.01** |
| Female | 0.27 | 0.10 | 0.44 | 0.10 | 0.00** |
| Previous mental health issues | 1.05 | 0.86 | 1.25 | 0.34 | 0.00*** |
| **Adjusted R^2^=0.15, n=853** |  |  |  |  |  |
| Step 3 |  |  |  |  |  |
| Perceive Risk of COVID-19 (per unit) | 0.02 | -0.01 | 0.04 | 0.03 | 0.22 |
| Female | 0.22 | 0.07 | 0.36 | 0.08 | 0.00** |
| Previous mental health issues | 0.45 | 0.28 | 0.62 | 0.15 | 0.00*** |
| Positive Mood (per unit) | -0.11 | -0.13 | -0.09 | -0.41 | 0.00*** |
| Perceived Loneliness (per unit)^b^ |  |  |  |  |  |
| Never | -0.28 | -0.49 | -0.07 | -0.08 | 0.01* |
| Occasionally | 0.24 | 0.05 | 0.43 | 0.08 | 0.01* |
| Some of the time | 0.34 | 0.14 | 0.54 | 0.11 | 0.00*** |
| Often/Always | 0.86 | 0.61 | 1.11 | 0.22 | 0.00*** |
| COVID-19 Worry^c^ |  |  |  |  |  |
| No worry | -0.11 | -0.27 | 0.05 | -0.04 | 0.19 |
| Much of time | 0.07 | -0.13 | 0.27 | 0.02 | 0.5 |
| Most of time | 0.31 | -0.06 | 0.68 | 0.04 | 0.1 |
| **Adjusted R^2^=0.46, n=853** | | |  |  |  |

*** *p*<0.001, ** *p*<0.01, * *p*<0.05

^a^ A square-root transformation was applied to the dependent variable.

^b^ Comparison reference group “I have hardly ever felt lonely over the past two weeks”.

^c^ Comparison reference group “I occasionally worry about getting COVID-19”.

**Table S5 Regression model showing associations between explanatory variables and anxiety scores**

|  | **B** | **95% CI Lower** | **95% CI Upper** | ***β*** | ***p*** |
| --- | --- | --- | --- | --- | --- |
| **GAD-7 (anxiety) Total Score^a^** |  |  |  |  |  |
| Step 1 |  |  |  |  |  |
| Perceived Risk of COVID-19 (per unit) | 0.08 | 0.05 | 0.12 | 0.16 | 0.00*** |
| **Adjusted R^2^=0.02, n=893** |  |  |  |  |  |
| Step 2 |  |  |  |  |  |
| Perceive Risk of COVID-19 (per unit) | 0.06 | 0.03 | 0.09 | 0.11 | 0.00*** |
| Female | 0.33 | 0.17 | 0.49 | 0.12 | 0.00*** |
| Previous mental health issues | 1.25 | 1.07 | 1.44 | 0.41 | 0.00*** |
| **Adjusted R^2^=0.22, n=853** |  |  |  |  |  |
| Step 3 |  |  |  |  |  |
| Perceive Risk of COVID-19 (per unit) | 0.03 | 0 | 0.05 | 0.05 | 0.03* |
| Female | 0.23 | 0.09 | 0.37 | 0.09 | 0.00** |
| Previous mental health issues | 0.71 | 0.54 | 0.87 | 0.23 | 0.00*** |
| Positive Mood (per unit) | -0.1 | -0.12 | -0.09 | -0.38 | 0.00*** |
| Perceived Loneliness (per unit)^b^ |  |  |  |  |  |
| Never | -0.14 | -0.35 | 0.07 | -0.04 | 0.2 |
| Occasionally | 0.25 | 0.07 | 0.44 | 0.08 | 0.01** |
| Some of the time | 0.31 | 0.11 | 0.51 | 0.1 | 0.00** |
| Often/Always | 0.6 | 0.35 | 0.85 | 0.15 | 0.00*** |
| COVID-19 Worry^c^ |  |  |  |  |  |
| No worry | -0.27 | -0.43 | -0.11 | -0.09 | 0.00** |
| Much of time | 0.3 | 0.1 | 0.5 | 0.08 | 0.00** |
| Most of time | 0.51 | 0.15 | 0.88 | 0.07 | 0.01** |
| **Adjusted R^2^=0.46, n=853** | | |  |  |  |

*** *p*<0.001, ** *p*<0.01, * *p*<0.05

^a^ A square-root transformation was applied to the dependent variable.

^b^ Comparison reference group “I have hardly ever felt lonely over the past two weeks”.

^c^ Comparison reference group “I occasionally worry about getting COVID-19”.

**Table S6 Regression model showing associations between explanatory variables and stress scores**

|  | **B** | **95% CI Lower** | **95% CI Upper** | ***β*** | ***p*** |
| --- | --- | --- | --- | --- | --- |
| **PSS-4 (stress) Total Score** |  |  |  |  |  |
| Step 1 |  |  |  |  |  |
| Perceived Risk of COVID-19 (per unit) | 0.14 | 0.06 | 0.22 | 0.12 | 0.00*** |
| **Adjusted R^2^=0.01, n=893** | | | | | |
| Step 2 |  |  |  |  |  |
| Perceive Risk of COVID-19 (per unit) | 0.09 | 0.01 | 0.17 | 0.08 | 0.02* |
| Female | 0.45 | 0.06 | 0.85 | 0.07 | 0.03* |
| Previous mental health issues | 2.00 | 1.54 | 2.45 | 0.29 | 0.00*** |
| **Adjusted R^2^=0.10, n=853** |  |  |  |  |  |
| Step 3 |  |  |  |  |  |
| Perceive Risk of COVID-19 (per unit) | 0.02 | -0.04 | 0.08 | 0.02 | 0.55 |
| Female | 0.44 | 0.1 | 0.77 | 0.07 | 0.01* |
| Previous mental health issues | 0.63 | 0.23 | 1.02 | 0.09 | 0.00** |
| Positive Mood (per unit) | -0.3 | -0.34 | -0.26 | -0.48 | 0.00*** |
| Perceived Loneliness (per unit)^a^ |  |  |  |  |  |
| Never | 0.09 | -0.41 | 0.59 | 0.01 | 0.71 |
| Occasionally | 0.48 | 0.04 | 0.93 | 0.07 | 0.03* |
| Some of the time | 0.69 | 0.21 | 1.17 | 0.1 | 0.00** |
| Often/Always | 1.4 | 0.81 | 2 | 0.16 | 0.00*** |
| COVID-19 Worry^b^ |  |  |  |  |  |
| No worry | 0.09 | -0.29 | 0.48 | 0.01 | 0.64 |
| Much of time | 0.67 | 0.19 | 1.15 | 0.08 | 0.01** |
| Most of time | 0.29 | -0.58 | 1.17 | 0.02 | 0.51 |
| **Adjusted R^2^=0.41, n=853** | | |  |  |  |

*** *p*<0.001, ** *p*<0.01, * *p*<0.05

^a^ Comparison reference group “I have hardly ever felt lonely over the past two weeks”.

^b^ Comparison reference group “I occasionally worry about getting COVID-19”.

Approach to Analyses

Participant characteristics and outcome variables (depression, anxiety and stress scores) were summarised using appropriate statistics and histograms and scatterplots were examined. We conducted independent sample t-tests to compare previously published normative values in people with similar age. Examination of histograms indicated both depression and anxiety scores deviated from a normal distribution, however transformations or non-parametric tests were not suitable for these comparisons as only summary statistics not individual level data were available for normative data. While t-tests are robust to deviations from normality especially when sample sizes are large, results of these specific tests should be interpreted with appropriate caution. We conducted univariable linear regression analyses with 95% confidence intervals to explore the association between perceived risk of COVID-19 and mental health outcomes (depression, anxiety, stress). We conducted subsequent multivariable regression models to explore the independent contributions of non-modifiable factors (gender and previous mental health issues) and modifiable explanatory factors (perceived risk of COVID-19, positive mood, perceived loneliness, worry about contracting COVID-19) to explaining variation in the mental health outcomes. The variable perceived loneliness was treated as categorical variable in all models, with “hardly ever” as the reference value as this was the most common response. The variable assessing COVID-19 worry was also treated as a categorical variable in all models, with “occasional worry” treated as the reference value as this was the most common response. Assumptions of linear regression (normality and homoscedasticity of residuals, linearity with continuous variables) and presence of outliers were assessed graphically. Multicollinearity was checked for all models using variance inflation factors (VIF) and found to have acceptable levels. Square root transformations were used for depression and anxiety scores to satisfy assumptions. Robustness of the models was examined by removing data points with large residuals (<-3 or >3) and comparing results to the original models. No substantive effect on interpretation was observed in all models hence these results are not presented.

Statistical analyses were performed using STATA (version 16).
